# Assessment of ageing using global mass-spectrometry based metabolomics: A cross-cohort longitudinal study in the UK and Ireland

**DOI:** 10.64898/2026.02.09.26345895

**Authors:** Chung-Ho E. Lau, Elena Chekmeneva, Rui Pinto, Aisling M. O’Halloran, Daniel K.H. Chu, Abbas Dehghan, Ioanna Tzoulaki, Paul Elliott, Rose Anne Kenny, Cathal McCrory, Oliver Robinson

## Abstract

**Introduction:** Understanding the links between metabolism, ageing and age-related phenotypes may clarify the role of ageing in disease onset and improve risk prediction.

**Methods:** We conducted a cross-cohort assessment of biological age using broad-spectrum LC-MS metabolomics in 2,295 participants, aged 20-89, from the UK Airwave study (N=960) and The Irish Longitudinal Study of Ageing (N=1,335).

**Results:** N_2_,N_2_-dimethylguanosine, C-glycosyltryptophan, bile acid glucuronides, and zeta-carotene were associated with chronological age, frailty, and mortality. We developed a metabolomic clock that was highly predictive of chronological age (r = 0.92) in test samples. Metabolomic age acceleration was strongly correlated between study visits (*r* > 0.6). Each standard deviation higher metabolomic age acceleration (∼5 years) was associated with 43% higher mortality risk, 27% higher risk of mild cognitive impairment, and 10% increased risk of a higher frailty score in fully adjusted models.

**Discussion:** Our metabolomic clock provides a reproducible marker of generalised age-related disease risk.

## Introduction

Ageing is a gradual pathophysiological process and the predominant risk factor for almost all common chronic diseases, including cancers, diabetes, neurodegeneration, and cardiovascular disease (CVD) ^1,2^. Addressing ageing physiology, either through lifestyle or therapeutic intervention, may help prevent or delay the onset or mitigate the severity of multiple chronic diseases. This is particularly pertinent today, given the growing disease burden resulting from an ageing population. Lifespan has been increasing faster than health span for the last two centuries, and prolonging disease- and disability-free years has become a focus of ageing research in recent years^3^. Notably, the rate of age-related functional decline varies among individuals, and may be modified by lifestyle, physical and mental health, and other social and environmental risk factors such as socio-economic status^4^. Inter-individual ageing variability may be captured through assessment of biological aging - a series of cellular and molecular changes that may precede disease onset ^5^.

High-throughput omics technology has extended our knowledge of cellular processes and molecular pathways related to ageing and expedited the development of a new generation of biological ageing clocks, enabling biological age to be measured at scale in large population-based studies ^6^. The metabolome has enormous potential for assessing ageing phenotypes, given that multiple ageing processes, including hallmarks such as intercellular communication and nutrient sensing, are intrinsically linked with metabolism, ^2,7,8^. A number of ageing clocks have been developed using metabolomics data ^9–11^, but so far, the most established metabolomics ageing models have been derived using Nightingale-Health nuclear magnetic resonance spectroscopy (NMR) data ^12,13^, which only measures the most abundant metabolites and samples a fraction of the metabolic pathways, missing key ageing processes.

Clocks based on broad coverage non-targeted liquid chromatography-mass spectrometry (LC-MS) metabolomics have demonstrated strong predictive performance for age, but a lack of metabolite annotation of model predictors has limited interpretability^9,14^. Furthermore, partly owing to the overreliance on non-standardised analytical methods, assessing cross-study applicability of metabolomic clocks remains a considerable challenge.

We aimed to perform a cross-cohort assessment of biological age through broad-spectrum LC-MS metabolomics, using both lipidomic and small molecule assays, following the open platform data processing protocols available at the National Phenome Centre ^15^. We analysed over 3,500 plasma samples collected from over 2,000 individuals ranging from 20 to 89 years old at baseline, from two nationwide cohorts in the UK and Ireland. Our approach combined untargeted metabolic feature discovery to expand coverage of age-related pathways, with metabolite identification and extraction for global metabolite profiling. Our objectives were to i) examine and compare metabolic associations with chronological age, frailty and mortality risk; ii) compute a metabolomic ageing clock to provide an overall assessment of metabolic ageing; and iii) investigate the relationship between accelerated metabolomic ageing with risk factors, mortality, and multiple ageing-related phenotypes including cognitive impairment.

## Methods

### Study Populations

The Airwave Health Monitoring Study (Airwave) is an occupational cohort study of participants from 28 UK police forces recruited from 2006 to 2015 ^16^. It was established to evaluate possible health risks associated with the use of TETRA, a digital communication system used by the police forces and other emergency services in Great Britain since 2001. The study involved a total of 28 (of 54) forces and enrolled 53,245 participants from 2004 to 2015, while 18,326 (40.2%) of these participants attended a follow-up visit on average 8.1 years later. Baseline and follow-up samples from 1,000 participants were selected for metabolomics analyses based on the availability of follow-up health data, with this sample being broadly representative of the whole Airwave cohort. Samples from 960 Airwave participants passed metabolomic QC and were included in the study analysis. Ethical approval was obtained from the National Health Service Multi-Site Research Ethics Committee (MREC/13/NW/0588).

The Irish Longitudinal Study of Aging (TILDA) is a prospective population-representative study of adults aged ≥50 years in Ireland with surveys conducted in two-year intervals since 2009. Participants were randomly selected based on their residential address, so that each residential address in Ireland had an equal probability of selection. Community-dwelling adults aged 50 years and over and their spouses (of any age) who were non-demented and able to provide informed consent were eligible to participate. In total 8,504 community-dwelling adults were enrolled in Wave 1 (2009-2011) ^17, 18^. A total of 1,400 Wave 1 samples and 500 Wave 3 repeat samples were selected for metabolomics analysis. Of the 1,400 Wave 1 participants, 400 had passed away by January 2022. Mortality cases are enriched in the sample selected to enable an independent matched case-control mortality analysis for model validation. Ethical approval was obtained from the Trinity College Dublin Faculty of Health Sciences Research Ethics Committee. Data in both cohorts were collected on sociodemographics, cognitive ability, lifestyle, and physical and mental health. A summary of participants included in the current study is provided in Figure S1.

### Metabolomic analyses and data processing

Broad-spectrum discovery metabolomics was performed on non-fasting blood plasma samples using ultra-performance LC-MS reversed-phase chromatography lipidome and small molecule profiling methods developed in-house at the National Phenome Centre, at Imperial College London ^15^. Specifically, analytes were separated using reversed-phase chromatography, and C_8_ and C_18_ columns were respectively utilised as the stationary phases for lipidomic and small molecule profiling, with full sample data acquired in both the electrospray positive (ESI+) and electrospray negative (ESI-) ionization modes, leading to four distinct datasets. Quality control (QC) samples, including pooled study reference, dilution series and long-term reference samples (LTR), were used to support the assessment of data reproducibility and linear dynamic ranges, and run order corrections between and within batches. Metabolite features with relative standard deviations of > 30% in the QC samples, or those with peak intensities exhibiting Pearson’s correlation coefficients < 0.7 with the dilution series, were subsequently excluded from further data analyses. The LC-MS metabolomics peak signal intensity data were initially processed using XCMS ^19^, producing 6,856 and 5,234 features, respectively, in Airwave and TILDA for untargeted discovery of age-related features. Following structural annotation of features through database search, tandem MS experiments, spectral interpretation, and comparison to authentic reference standards when available, a final set of 569 relatively quantified metabolites was extracted for statistical analysis, using the *peakPantheR* package developed at the National Phenome Centre ^20^. Signals from the 569 relatively quantified metabolites were additionally scaled to the corresponding signals from LTR samples (commercially sourced plasma sample maintained by the National Phenome Centre and utilized as an independent sample reference for relating measurements across multiple profiling studies) prior to statistical analysis to minimise any analytical batch effects between cohorts. A detailed description of analytical protocols and quality control procedures has been published previously ^15^, while details on metabolite annotation can be found in Table S1 and confidence in annotations were assigned according to the Metabolomics Standards Initiative ^21^. To compare the molecular coverage of the ageing metabolome, in addition to those analysed at the National Phenome Centre, we included a subset of Airwave samples (N = 148) also analysed using the commercially available Metabolon (Durham, NC, USA) and Nightingale Health metabolomics platforms ^22-24^.

### Health outcomes and covariates

In both cohorts, hypertension was defined based on medical diagnosis, use of antihypertensive medications, systolic blood pressure >= 140 mmHg, or diastolic blood pressure >= 90 mmHg. Diabetes was defined based on medical diagnosis, use of diabetic medications, or a HbA1c level >= 6.5% (48mmol/mol). A BMI >= 30 was defined as obesity. For smoking, participants were categorised into never-smokers, former smokers, and current smokers. Participants were categorised based on their highest level of educational attainment (primary, secondary, or tertiary). Physical activities were determined using the International Physical Activity Questionnaire, with participants who achieved < 600 MET-mins/week being considered physically inactive. Depression risk was assessed in Airwave using PHQ-9 (Patient Health Questionnaire-9), a nine-item questionnaire, and individuals are considered at risk if they scored over 9/27. In TILDA, depression risk was assessed using the Center for Epidemiologic Studies Depression Scale, with those scoring >= 16 out of 60 considered at risk. Individuals in Airwave were considered heavy alcohol users if they consumed more than the UK’s recommended guidelines of 14 or 21 units/week for women and men, respectively. In TILDA, heavy alcohol use was indicated if participants scored one or more in the four-item CAGE questionnaire. Household income was analysed as a binary variable, with households earning < £26,000 in Airwave or those earning < €15,000 per annum in TILDA considered as low income. Antihypertensive and diabetic medications data were based on self-report.

Within TILDA, mortality data were extracted via linkage with the Irish national death registry until the end of January 2022, when participants were followed up for 11 years. Frailty was assessed using the composite Fried Frailty Index, which consists of five different domains: exhaustion, unexplained weight loss, weakness, slowness, and low physical activity ^25^. Fried Frailty Index was scored from 0 to 5, with the presence of each component contributing one unit to the total score. Timed-up-and-Go (TUG) was used to assess mobility, where participants were asked to stand from a seated position, walk 3 meters at their usual pace, turn around, walk back to the chair, and sit down again. TUG was measured using a chair with armrests and a seat height of 46 cm at Wave 1, ^26–28^. Hand grip strength was measured using a hydraulic hand dynamometer during the health assessment at Wave 1 ^27^. Cognitive assessments in TILDA used the Mini-Mental State Examination (MMSE) and Montreal Cognitive Assessment (MoCA), both well-established screening tools. These were administered by trained nursing staff to evaluate global cognitive performance. The MMSE includes measures of orientation, registration, attention, calculation, recall, and language.

MoCA additionally assesses executive function, abstraction, and visuospatial ability ^29^. MMSE and MoCA are both scored out of 30. MoCA is known to be more sensitive to mild cognitive deficits than the MMSE, and we used a MoCA score of less than 24 as a conservative cutoff of Mild Cognitive Impairment (MCI) ^30^. The Mini-Mental State Examination (MMSE) measurements were obtained at baseline (W1) and up to four subsequent study visits (W2-W5); MMSE data were available in 1,331 participants, and five repeated measurements were available in 827 participants. The choice reaction time test is a computer-based program to assess concentration and processing speed; motor response time (ms) was recorded as participants responded to stimuli shown on-screen ^31^. Extrinsic epigenetic age accelerations (EEAA) were calculated for Horvath, Hannum, GrimAge, PhenoAge clocks, which were estimated using the Infinium Methylation EPIC BeadChip data collected from whole blood samples with adjustment for chronological age. Leukocyte telomere length was calculated as the mean telomere to single copy gene (T/S) ratio as determined by monochrome multiplex quantitative PCR using DNA extracted directly from the blood samples taken during the health assessment. Non-fasting samples were used for measuring clinical blood markers, including C-Reactive Protein (CRP), HbA1c, total cholesterol, and HDL. Renal markers creatinine and Cystatin were measured from frozen plasma ^32^. Cardiovascular diseases (CVD) conditions in TILDA were based on self-report CAPI questionnaire data.

### Statistical Analyses

All statistical analyses were performed using packages available in R 4.5.2.

### Metabolomic-wide association studies

Metabolite levels were natural-log transformed, mean-centered, and unit-variance scaled before linear regression analysis and were treated as the dependent variables. To compare associations with chronological age in both cohorts, metabolite associations were tested using multiple linear regressions, adjusted for sex and BMI, and Bonferroni’s method was used to account for multiple testing.

Within the TILDA population only, we compared metabolite associations for chronological age, Fried frailty index (negative binomial regression using *glm.nb* function from the *MASS* package) and all-cause mortality (Cox proportional hazards regression). In this analysis, chronological age associations were adjusted only for sex, as a direct association between BMI and age was observed only in Airwave but not in TILDA (Figure S2). To better assess the direct associations with the metabolome, Fried frailty index analyses were adjusted for age, sex, hypertension, diabetes, statin use, number of medications, depression, education, smoking, and alcohol use. Mortality analyses were additionally adjusted for BMI and physical activity. We excluded BMI and physical activity as covariates in the analyses involving the frailty index, as they are closely related to elements of the Fried frailty index. For comparison of metabolite associations with these metrics in TILDA, significant metabolite associations after adjusting for false discovery rate at 5% were reported.

### Bootstrap aggregate LASSO model of chronological age

To improve the overall model’s accuracy and robustness, applying a similar methodology as recently used for proteomic clock development, ^33^ we used bootstrap aggregation for model training in constructing the metabolomic predictor of chronological age. LASSO models were trained using the *glmnet* R package, and our training data consist of an equal number of 683 baseline samples from each of the two cohorts (N = 1,366, Table 1), including 569 metabolites as model input, where sex was included as an unpenalized model predictor. To avoid overfitting λ_1SE_ was used as the model λ and was determined through five-fold cross-validation using the *cv.glmnet* function. We resampled with replacement to generate 100 bootstrap samples of the training data, resulting in 100 bootstrap models created, and their mean predicted age was used for calculating metabolomic age gaps. Additionally, to assess the relative utility of lipids and small molecules for modelling chronological age, we separately constructed multivariable age prediction models using metabolites only detected in either the lipid profile (NPC-lipid) or the small molecule profile (NPC-small), which included 387 and 180 metabolites as model inputs, respectively.

**Table 1.** Characteristics and demographics of study sample population.

| Baseline | Training sample |  | Validation |  |  | Total |  |
| --- | --- | --- | --- | --- | --- | --- | --- |
|  | AIRWAVE<br>(N=683) | TILDA<br>(N=683) | AIRWAVE<br>(N=277) | TILDA<br>(N=326)<br>Control | TILDA<br>(N=326)<br>Deaths | AIRWAVE<br>(N=960) | TILDA<br>(N=1335) |
| **Age** |  |  |  |  |  |  |  |
| Mean (SD) | 41.7 (9.58) | 60.6 (9.09) | 41.2 (9.02) | 68.9 (7.92) | 69.2 (7.98) | 41.6 (9.42) | 64.7 (9.53) |
| Median [Min, Max] | 42.0 [20.0, 68.0] | 58.0 [50.0, 89.0] | 42.0 [20.0, 67.0] | 70.0 [50.0, 86.0] | 70.0 [50.0, 85.0] | 42.0 [20.0, 68.0] | 64.0 [50.0, 89.0] |
| **Sex** |  |  |  |  |  |  |  |
| Female | 257 (37.6%) | 410 (60.0%) | 105 (37.9%) | 142 (43.6%) | 142 (43.6%) | 362 (37.7%) | 694 (52.0%) |
| Male | 426 (62.4%) | 273 (40.0%) | 172 (62.1%) | 184 (56.4%) | 184 (56.4%) | 598 (62.3%) | 641 (48.0%) |
| **BMI** |  |  |  |  |  |  |  |
| Mean (SD) | 26.5 (4.09) | 29.1 (6.13) | 26.5 (3.66) | 28.4 (4.15) | 28.2 (4.33) | 26.5 (3.97) | 28.7 (5.30) |
| Median [Min, Max] | 26.1 [17.9, 48.2] | 28.0 [16.5, 65.2] | 26.2 [18.3, 39.8] | 28.1 [18.5, 43.3] | 28.1 [19.0, 41.9] | 26.2 [17.9, 48.2] | 28.1 [16.5, 65.2] |
| Missing | 0 (0%) | 1 (0.1%) | 1 (0.4%) | 0 (0%) | 0 (0%) | 1 (0.1%) | 1 (0.1%) |
| **Hypertension** |  |  |  |  |  |  |  |
| No | 470 (68.8%) | 278 (40.7%) | 203 (73.3%) | 92 (28.2%) | 75 (23.0%) | 673 (70.1%) | 445 (33.3%) |
| Yes | 213 (31.2%) | 404 (59.2%) | 74 (26.7%) | 234 (71.8%) | 250 (76.7%) | 287 (29.9%) | 888 (66.5%) |
| Missing | 0 (0%) | 1 (0.1%) | 0 (0%) | 0 (0%) | 1 (0.3%) | 0 (0%) | 2 (0.1%) |
| **Diabetes** |  |  |  |  |  |  |  |
| No | 660 (96.6%) | 615 (90.0%) | 268 (96.8%) | 286 (87.7%) | 275 (84.4%) | 928 (96.7%) | 1176 (88.1%) |
| Yes | 23 (3.4%) | 58 (8.5%) | 9 (3.2%) | 28 (8.6%) | 41 (12.6%) | 32 (3.3%) | 127 (9.5%) |
| Missing | 0 (0%) | 10 (1.5%) | 0 (0%) | 12 (3.7%) | 10 (3.1%) | 0 (0%) | 32 (2.4%) |
| **Statin Use** |  |  |  |  |  |  |  |
| No | 653 (95.6%) | 500 (73.2%) | 267 (96.4%) | 211 (64.7%) | 197 (60.4%) | 920 (95.8%) | 908 (68.0%) |
| Yes | 30 (4.4%) | 183 (26.8%) | 10 (3.6%) | 115 (35.3%) | 129 (39.6%) | 40 (4.2%) | 427 (32.0%) |
| **Num Medication Use** |  |  |  |  |  |  |  |
| Mean (SD) | - | 2.41 (2.67) | - | 3.15 (2.76) | 3.98 (3.08) | - | 2.97 (2.87) |
| Missing | 683 (100%) | 2 (0.3%) | 277 (100%) | 1 (0.3%) | 6 (1.8%) | 960 (100%) | 9 (0.7%) |
| **Depression** |  |  |  |  |  |  |  |
| No-Minimal symptoms | 631 (92.4%) | 617 (90.3%) | 259 (93.5%) | 309 (94.8%) | 275 (84.4%) | 890 (92.7%) | 1201 (90.0%) |
| At risk | 52 (7.6%) | 56 (8.2%) | 18 (6.5%) | 17 (5.2%) | 47 (14.4%) | 70 (7.3%) | 120 (9.0%) |
| Missing | 0 (0%) | 10 (1.5%) | 0 (0%) | 0 (0%) | 4 (1.2%) | 0 (0%) | 14 (1.0%) |
| **Education** |  |  |  |  |  |  |  |
| Tertiary | 203 (29.7%) | 256 (37.5%) | 83 (30.0%) | 129 (39.6%) | 74 (22.7%) | 286 (29.8%) | 459 (34.4%) |
| Secondary | 454 (66.5%) | 280 (41.0%) | 185 (66.8%) | 105 (32.2%) | 120 (36.8%) | 639 (66.6%) | 505 (37.8%) |
| Primary | 26 (3.8%) | 145 (21.2%) | 9 (3.2%) | 91 (27.9%) | 131 (40.2%) | 35 (3.6%) | 367 (27.5%) |
| Missing | 0 (0%) | 2 (0.3%) | 0 (0%) | 1 (0.3%) | 1 (0.3%) | 0 (0%) | 4 (0.3%) |
| **Smoking** |  |  |  |  |  |  |  |
| Never | 453 (66.3%) | 288 (42.2%) | 199 (71.8%) | 147 (45.1%) | 109 (33.4%) | 652 (67.9%) | 544 (40.7%) |
| Past | 181 (26.5%) | 282 (41.3%) | 62 (22.4%) | 151 (46.3%) | 125 (38.3%) | 243 (25.3%) | 558 (41.8%) |
| Current | 49 (7.2%) | 113 (16.5%) | 16 (5.8%) | 28 (8.6%) | 92 (28.2%) | 65 (6.8%) | 233 (17.5%) |
| **Alcohol Use** |  |  |  |  |  |  |  |
| Less than Heavy | 628 (91.9%) | 473 (69.3%) | 262 (94.6%) | 241 (73.9%) | 220 (67.5%) | 890 (92.7%) | 934 (70.0%) |
| Heavy | 55 (8.1%) | 154 (22.5%) | 15 (5.4%) | 61 (18.7%) | 69 (21.2%) | 70 (7.3%) | 284 (21.3%) |
| Missing | 0 (0%) | 56 (8.2%) | 0 (0%) | 24 (7.4%) | 37 (11.3%) | 0 (0%) | 117 (8.8%) |
| **Physical activity** |  |  |  |  |  |  |  |
| Active | 395 (57.8%) | 518 (75.8%) | 162 (58.5%) | 266 (81.6%) | 205 (62.9%) | 557 (58.0%) | 989 (74.1%) |
| Inactive | 8 (1.2%) | 156 (22.8%) | 4 (1.4%) | 58 (17.8%) | 121 (37.1%) | 12 (1.3%) | 335 (25.1%) |
| Missing | 280 (41.0%) | 9 (1.3%) | 111 (40.1%) | 2 (0.6%) | 0 (0%) | 391 (40.7%) | 11 (0.8%) |
| **Household income** |  |  |  |  |  |  |  |
| High | 644 (94.3%) | 435 (63.7%) | 248 (89.5%) | 197 (60.4%) | 172 (52.8%) | 892 (92.9%) | 804 (60.2%) |
| Low | 39 (5.7%) | 99 (14.5%) | 29 (10.5%) | 51 (15.6%) | 85 (26.1%) | 68 (7.1%) | 235 (17.6%) |
| Missing | 0 (0%) | 149 (21.8%) | 0 (0%) | 78 (23.9%) | 69 (21.2%) | 0 (0%) | 296 (22.2%) |
| **Died during follow-up** |  |  |  |  |  |  |  |
| No | - | 590 (86.4%) | - | 326 (100%) | 0 | - | 916 (68.6%) |
| Yes | - | 93 (13.6%) | - | 0 | 326 (100%) | - | 419 (31.4%) |
| **Repeat visits** |  |  |  |  |  | **(N = 914)** | **(N = 477)** |
| **Age** |  |  |  |  |  |  |  |
| Mean (SD) |  |  |  |  |  | 48.6 (9.38) | 67.4 (8.94) |
| Median [Min, Max] |  |  |  |  |  | 49.0 [27.0, 74.0] | 66.0 [54.0, 89.0] |
| **Sex** |  |  |  |  |  |  |  |
| Female |  |  |  |  |  | 348 (38.1%) | 261 (54.7%) |
| Male |  |  |  |  |  | 566 (61.9%) | 216 (45.3%) |
| **BMI** |  |  |  |  |  |  |  |
| Mean (SD) |  |  |  |  |  | 27.0 (4.11) | 28.9 (5.43) |
| Median [Min, Max] |  |  |  |  |  | 26.6 [18.3, 49.5] | 28.4 [16.8, 63.1] |
| Missing |  |  |  |  |  | 0 (0%) | 15 (3.1%) |

### Analysis of metabolomic age gap with aging phenotypes

Age gaps were calculated separately per cohort and study timepoints by obtaining the linear regression residuals of the predicted metabolomic age from chronological age. Associations between age gap and all-cause mortality were assessed in a subset of TILDA mortality case-control samples excluded from model training. These controls and mortality case samples were age, sex and BMI matched (N *case* = 326) and survival analyses were performed with three different set of covariate adjustments, with Model 1adjusted for age and sex, Model 2 additionally adjusted for potential biological factors (BMI, hypertension, diabetes, statin use, number of medication use), and Model 3 additionally adjusted for common risk factors (depression, education, smoking, alcohol use and physical activity). Hazard ratios were estimated per SD change and per year change in age gaps. Area under the ROC curves (AUC) were estimated for 10-year all-cause mortality risk using the validation samples in the TIDLA cohort (N *case* = 326) using the *pROC* package. We compared the AUC of two models, clinical risk factor model (age, sex, BMI, alcohol use, smoking, diabetes, hypertension, education, physical inactivity, depression, statin use, medication use) against a model additionally including metabolomic age gap as covariate, and test for significance using the bootstrap method available from the *roc.test* function.

Associations with Fried Frailty Index and MCI were assessed using negative binomial regression and logistic regression with the same covariate adjustment sets used for mortality, except for the exclusion of BMI and physical activity in the frailty analysis. Mixed effect models, adjusting for baseline age, sex, time elapsed (fixed effect), and subject (random effect), were used to assess associations between the metabolomic age gap, estimated at baseline, and repeated cognitive performance in the MiniLMental State Examination (MMSE).

For analyses of age gaps with disease risk factors and ageing phenotypes, age gaps were standardised and treated as dependent variables in linear regression models, and significant associations with nominal P < 0.05 were reported. Lower disease risk groups were used as model reference groups in the risk factor analyses; current smokers were compared with non-smokers (reference group), participants with primary education as their highest qualification were compared to those with tertiary education (reference group), and heavy drinkers were compared against all other groups (reference group) as the variable was binarized. Regression analyses in this study were not adjusted for ethnicity since TILDA was ubiquitously White Irish, and approximately 99% of the sample Airwave cohort population were White European.

## Results

### Study population

We were particularly interested in identifying associations between metabolite and population ageing that may be generalisable across different study populations, thus in this study we have included 3,686 metabolomics samples from 2,295 individuals from two, predominantly of White European (∼ 99%), but otherwise demographically distinctive cohort studies in Airwave (n _individuals_ = 960) and TILDA (n _individuals_ = 1,335). Also, with a median age of 42 years (range: 20 – 68 years) in Airwave and a median age of 58 years (range: 50– 89 years) in TILDA at baseline, the sample distributions of chronological age of the two populations are highly complementary, permitting us to model a comprehensive spectrum of adult lifespan in the study analyses.

Given the TILDA cohort is an older adult population compared to the Airwave cohort, a higher proportion of individuals were affected by chronic conditions (obesity, hypertension, or diabetes), were current smokers, or were physically inactive than in the Airwave Study. Detailed sample characteristics of the study samples are reported in Table 1 and further cohort information is available in the Methods.

### Identifying signatures of chronological age with non-targeted metabolomics

To capture the full range of metabolic variation linked to population ageing while minimizing redundancy, age-related metabolite discovery was performed using a two-step approach. (Figure 1). As the metabolomics data were derived from four non-targeted LC-MS profiling assays (lipid profile ^ESI+^, lipid profile ^ESI-^, RPC small molecule ^ESI+^, RPC small molecule ^ESI-^), we first conducted preliminary univariable regression analyses against chronological age, adjusted for sex and BMI, using the combined unannotated feature variable data (Airwave N _features_ = 6,856 and TILDA N _features_ =5,234), separately for each of the two sample populations (Figure S3). This allowed us to screen the metabolomic dataset for information relating to chronological age and expand on our pre-existing assay compound target database by additionally annotating the most consistent and strongest age-related features (Table S1, Table S2). The 67 additionally selected and annotated age-related features, which were predominantly small molecules, were then combined with the pre-existing compound database for targeted peak signal extraction to provide a more comprehensive set of metabolite signals for subsequent analysis. Of the 569 metabolites examined, 246 and 253 age associations were found in Airwave and TILDA after Bonferroni multiple testing correction (Figure 2A, Table S2), of which 83 were concordant across the two cohorts, including 63 small molecules. The metabolites included a large number of steroids (negatively correlated with age), glucuronides and sulphated compounds of steroids. Other notable, strongest chronological age associations consistent across both cohorts included sialyllactose, pentose sugar, methylascorbic acid, 3-carboxy-4-methyl-5-pentyl-2-furanpropanoic acid (3-CMPFP), benzoylcarnitine, phosphatidylcholine PC(18:1/22:6), N-acetylglutamic acid, N-acetyltyrosine, hippuric acid, and C-glycosyltryptophan (all positive associations with P < 10^-10^, Figure 2A). Whilst free fatty acids and acylcarnitine levels generally increased with chronological age (Figure 3A), in keeping with our earlier results, our findings indicate that the strongest chronological age associations are more likely to be derived from small molecules and not from lipid molecules. Whilst we were not able to confirm their structures, two further unknown molecules, annotated as C_9_H_17_NO_3_ (detected in RPC small molecule as [M+H]^+^ *m/z* 188.1285, Figure S7) and C_14_H_27_O_7_P (detected in RPC small molecule as [M-H]- *m/z* 337.142), were found respectively, strongly positively and negatively associated with chronological age. The unknown molecule C_9_H_17_NO_3_ (Figure S7) has already been reported as positively correlated with chronological age in a metabolomics study of biological processes of ageing in twins ^34^.

**Figure 1.**
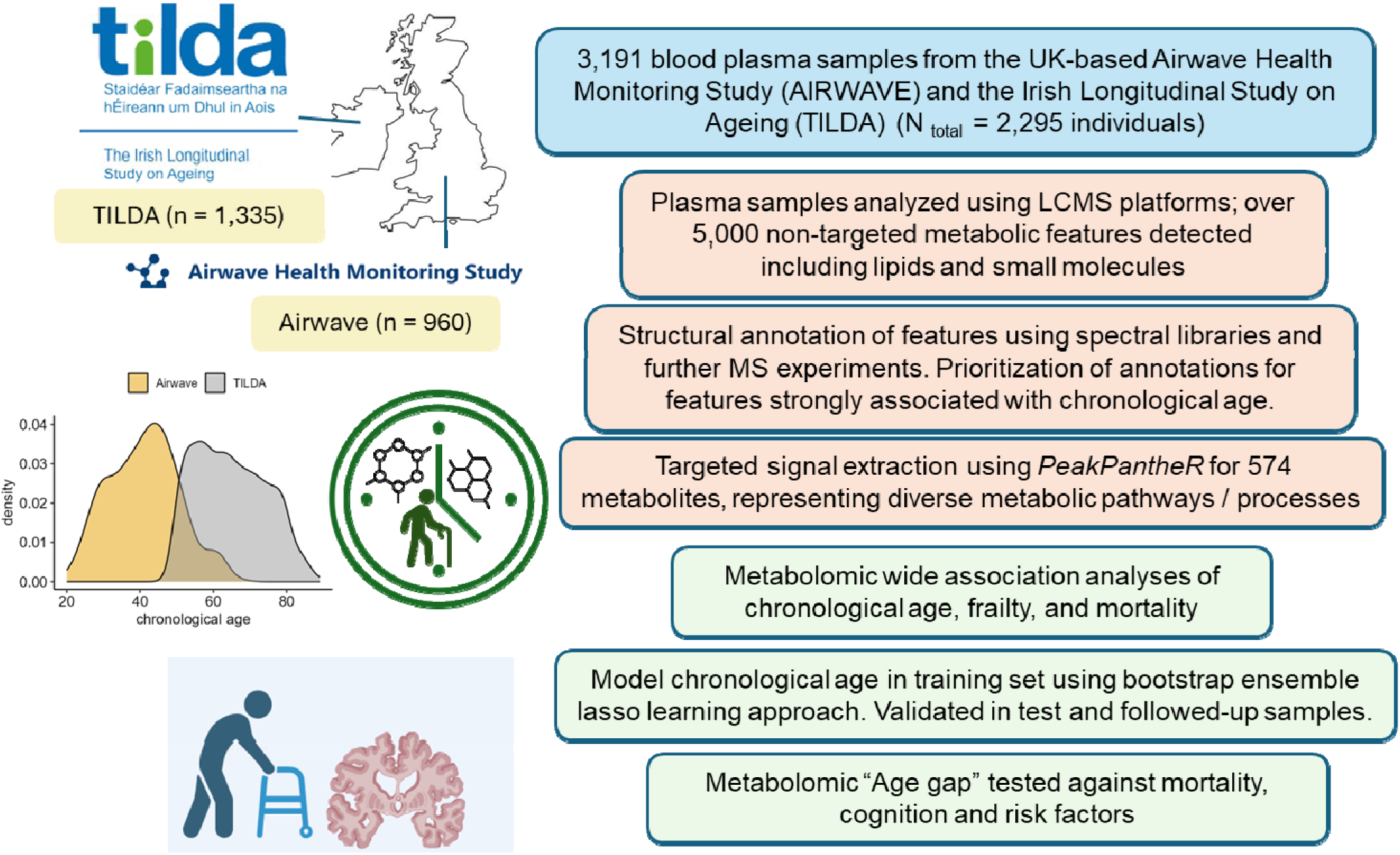
Study Workflow

**Figure 2.**
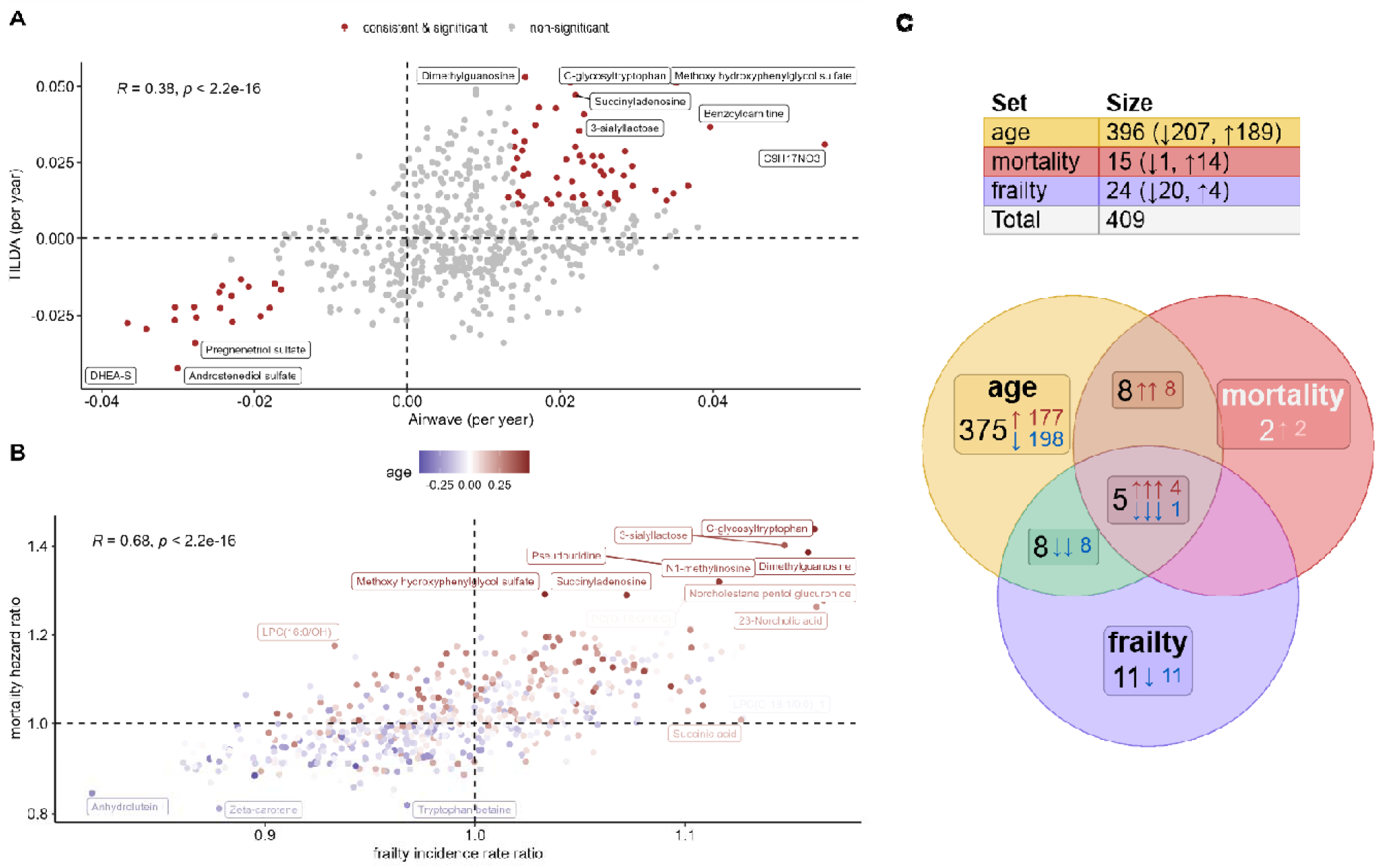
Metabolic associations of chronological age, frailty and mortality. A) Metabolomic wide association study of chronological age analyses were compared between the two study cohorts, adjusting for sex and BMI B) Comparison of Metabolomic wide association studies of chronological age, Fried frailty index (negative binomial regression) and all-cause mortality (Cox proportional hazards regression) were compared in the TILDA wave 1 samples. Chronological age analyses were adjusted for sex. Fried frailty index analyses were adjusted for age, sex, hypertension, diabetes, statin use, number of medications used, depression, education, smoking, alcohol use, and mortality analyses were additionally adjusted for BMI and physical activity. C) Venn diagram shows the overlap of metabolic associations. There are 419 death events recorded (follow-up of 11 years) and information of frailty index was available for 1,296 participants, and 494 participants have a frailty score of 1 or higher.

**Figure 3.**
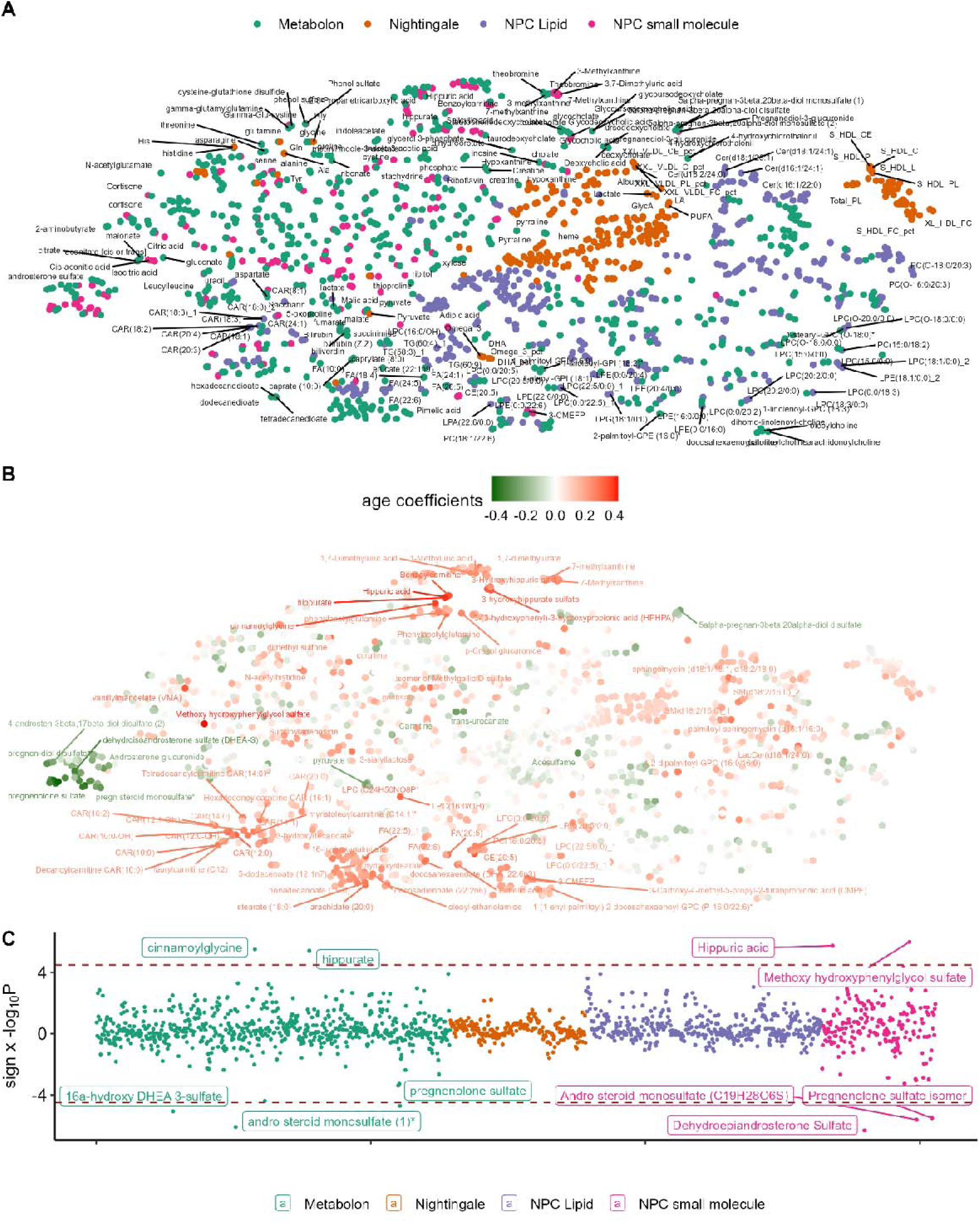
Cross-platform comparison of chronological age associations of Airwave sample subset analysed using the Metabolon and Nightingale metabolomics. t-SNE map of the combined 1,534 metabolic variables from across our in-house Metabolon and Nightingale metabolomics platforms, coloured A) by platform, and B) by standardized coefficients of chronological age associations adjusted for sex and BMI. C) Signed Manhattan plot comparing the statistical significance of age associations detected across the metabolomics platforms. N = 148.

The comparison of chronological age associations obtained in our two assays to those measured by Metabolon and Nightingale Health in a subset of Airwave participants (N = 148) revealed that our lipid profiling and small molecule profiling assays provided comparable and broad metabolite coverage relevant to population ageing (Figure 3A, Figure 3C). The gut microbial co-metabolite, hippurate, and steroid sulphates (pregnenolone sulphate, androsteroid monosulphate, DHEA-S metabolites, Figure 3B) were respectively the most robust positive and negative chronological age associations in both Metabolon and our assay platforms. Nightingale Health, which contains predominantly measurements from lipoproteins, demonstrated relatively weak associations with chronological age in this subset of Airwave samples.

### Metabolite associations with chronological age, frailty and mortality in TILDA

Next, we examined if metabolite associations of chronological age were related to mortality and frailty – two key metrics of functional ageing of clinical relevance - making use of the all-cause mortality data (N _case_ = 419, follow-up time ∼ 11 years) and the Fried frailty index (N = 1,296), available in the older TILDA study population In adjusted analyses, after correction for false discovery rate, 396 out of 569 metabolites (70%) tested were associated with chronological age in TILDA, and only 15 (14↑, 1↓) and 24 (20 ↓, 4↑) metabolites were associated with all-cause mortality and frailty in Cox proportional hazard regression and negative binomial regression analyses respectively (see Methods). Six of those were consistently associated with chronological age, frailty and mortality in the study sample (Figure 2C, Table S3). Four were positive associations: N_2_,N_2_-dimethylguanosine, C-glycosyltryptophan, norcholestane pentol glucuronide, and norcholic acid; and the only metabolite found negatively associated with all three ageing parameters was a carotenoid (zeta-carotene), which has strong antioxidant properties and is naturally produced in plants. Notably, other metabolites negatively associated with frailty were polyunsaturated fatty acid (PUFA) containing triglycerides (TG), luteine (a carotenoid) and retinol (vitamin A1); whilst sialyllactose, 3-methoxy-4-hydroxyphenylglycol sulfate (MHPG-S), sucrose, pseudouridine, o-sulfotyrosine, N_1_-methylinosine, succinyladenosine, a lysophosphatidylcholine (C24H50NO8P), pyroglutamyltaurine were found directly associated with both age and mortality. Notably, the estimates of metabolite associations with Fried frailty and mortality (Pearson’s *r* = 0.68, Figure 2B) were more strongly correlated than metabolite associations between age and mortality (*r* = 0.51), and age and frailty (*r* = 0.35).

### Metabolomic clock development

For metabolomic clock development (Methods), the training set contained an equal number of 683 baseline observations from each cohort, broadly representative of the wider cohort samples, and included 93 participants from TILDA who died during follow-up (Table 1). A total of 227 and 652 baseline samples and 914 and 477 follow-up samples, respectively, from Airwave and TILDA, were reserved as a testing set to evaluate the model’s predictive performance. Our predicted metabolomic age was strongly correlated with chronological age (*r* = 0.92, P < 2 x 10-^16^) with a mean absolute error (MAE) of 4.8 years in our independent test samples (Figure 4A). Furthermore, we were able to further evaluate how our model performed longitudinally in the same individuals (Figure 4B), and found the follow-up samples to have an average increase in predicted age of 4.2 yrs (SD: 3 ys) over a mean follow-up time of 7 yrs in Airwave, and 2.0 yrs increase in predicted age (SD: 4 yrs) over a mean follow-up time of 4 yrs in TILDA. Additionally, our data also suggest that metabolomic age gaps – the model residualised difference of predicted and chronological age, estimated at baseline and subsequent visit samples - were highly consistent (Pearson’s *r* = 0.66, P < 2 x 10-^16^, Figure 4C), indicating the relative stability of our metabolomics-derived biological ageing metric in the population setting. The most important contributors to the metabolomic clock (Figure 4D and Table S4) included a diverse range of metabolite classes, including steroid sulphates and glucuronides, acyl-carnitines, gut microbial metabolites, vitamins (e.g. niacinamide), peptides and neurotransmission-related metabolites (e.g. kynurenine, MHPG-S).

**Figure 4.**
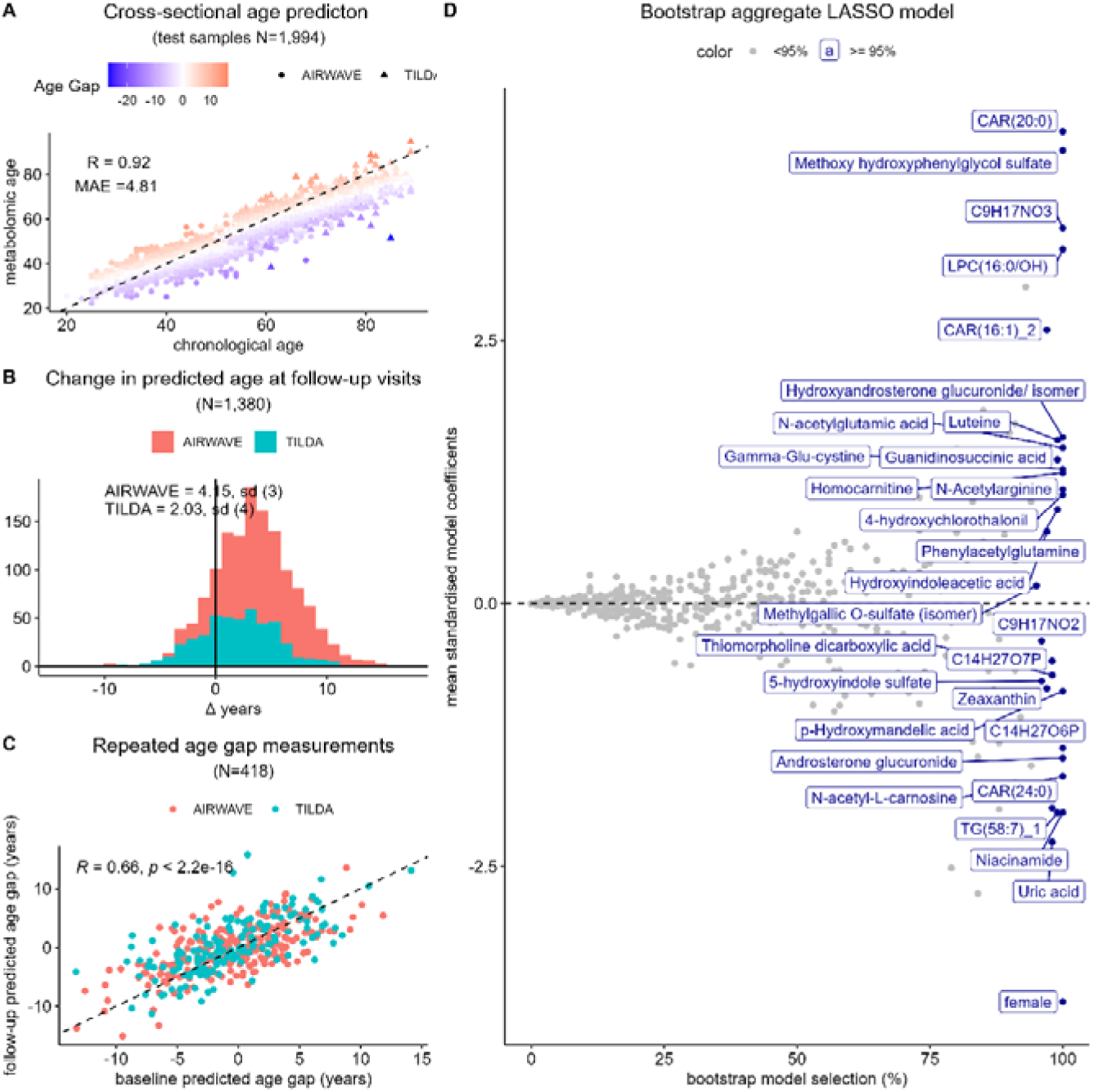
Bootstrap aggregate LASSO model of chronological age. A) Prediction performance in the test samples coloured by sample estimated metabolomic age gap; B) Longitudinal changes in predicted age in the followed-up samples; repeated measurements were taken after a 4-year interval in TILDA and after an average of 7 years (IQR:6-8 years) in Airwave C) Repeated age gaps measurements across the two study timepoints. D) 4 Bootstrap aggregate LASSO model predictor selection frequency, male and female were coded as 0 and 1 and were unpenalized in the LASSO models.

To better understand the relative utility of lipids and small molecules for modelling chronological age, we separately constructed age prediction models using only metabolites detected in either the lipid profile (NPC-lipid) or the small molecule profile (NPC-small) acquisition modes, and consistently found that small molecules had higher predictive performance in both cross-sectional (NPC-small MAE: 5.3 yrs vs NPC-lipid MAE: 5.8 yrs) and longitudinal settings in our cohort data (Figure S4 and Figure S5).

### Associations of metabolomic age gap with mortality, frailty and cognitive performance

To provide clinical validation of the metabolomic age gap as a metric of biological ageing, we examined its association with all-cause mortality in a subset involving an equal number of TILDA mortality case and control samples that had not previously been used for model training, matched for age, sex, and BMI (N _case/controls_ = 326). For our main metabolomic age assessment, the hazard ratios (HR) per standard deviation in age gap (equivalent to 4.8 years in age gap) for Model 1 were estimated to be 1.43 (CI:1.28-1.60, P = 2 x 10^-10^) for Model 1, 1.37 (CI:1.22 −1.54, P = 8 x 10^-8^) for Model 2 and 1.41 (CI:1.24 −1.60, P = 1 x 10^-7^) for Model 3 (see Methods for Model descriptions). For the NPC-main model, the hazard ratios per metabolomic age gap in years were estimated to be 1.07 in both Model 1 and Model 3 (Figure S6). As with the case for chronological age prediction, the NPC-lipid model showed weaker associations (P = 0.03 in Model 3) with mortality than the NPC-small molecule model (P = 2 x 10^-8^ in Model 3) (Figure 5A). Adding metabolomic age to clinical risk factors significantly improved predictive performance of area under the ROC curve (AUC) for 10-year all-cause mortality in the TILDA study validation samples (AUC-clinical + metabolomic age: 0.74, AUC-clinical: 0.71) (Figure 5B)

**Figure 5.**
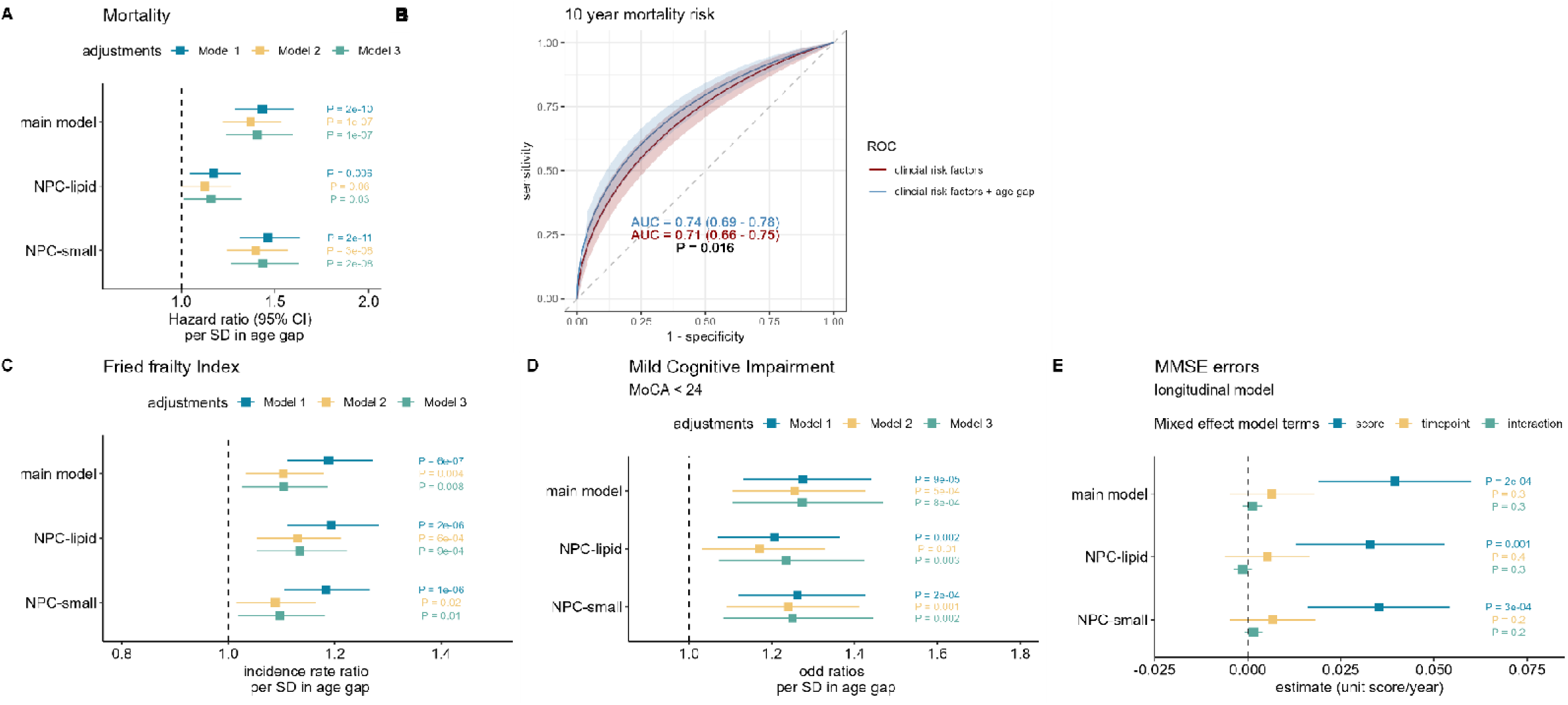
Metabolomic age gap and ageing phenotypes in TILDA. Model 1 adjusted for age and sex, Model 2 additionally adjusted BMI, hypertension, diabetes, statin use, number of medications used, Model 3 additionally adjusted for depression, education, smoking, alcohol use and physical activity. For analysis of frailty BMI and physical activity were excluded from adjustment sets. A) Associations between age gap and all-cause mortality, in controls and mortality case samples matched 1:1 for age, sex and BMI (N case = 326) excluded from model training. B) Receiver operating characteristic curve of 10 year mortality risk in TIDLA, comparing the clinical risk model (age, sex, BMI, alcohol use, smoking, diabetes, hypertension, education, physical inactivity, depression, statin use, medication use), and the clinical risk and metabolomic age gap model in the validation sample set. C) Associations between age gap and Fried frailty index in all TILDA samples. D) Associations between age gap and mild cognitive impairment, assessed through the use of Montreal Cognitive Assessment (MoCA) < 24. E) Assessment of baseline age gap (Wave 1) on repeated measurement of cognitive performance in Mini-Mental State Examination (Waves 1,2,3,4, and 5) using a linear mixed effect model, adjusted for age and sex.

We further assessed age gap associations with frailty and cognition, using a similar adjustment strategy, within the whole TILDA population. Although associations with the Fried frailty index were attenuated upon adjustment for clinical factors, all three metabolomic age gap models remained associated with a higher frailty index score in Model 3 (P < 0.05, Figure 5C), with the main metabolomic age gap assessment associated with 10% higher risks of frailty (incidence rate ratio for Fried frailty index: 1.10; 95% CI:1.03-1.19). We found that associations of the metabolomic age gap and MCI (MoCA <24) were broadly similar across the three covariate adjustment models tested, as each standard deviation increase in our main age gap model corresponded to approximately 25% higher risk of MCI (P ≤ 0.001, odds ratios: 1.25-1.27, CI: 1.10-1.47, Figure 5D). Additionally, we assessed the association between baseline age gap (Wave 1) on repeated measurements of cognitive performance in Mini-Mental State Examination (MMSE) using a linear mixed effect model (Figure 5E).

Higher age gaps at baseline were consistently associated with more frequent errors in the repeated MMSE assessments across the three age gap models tested (estimate: 0.04 errors per year in age gap; 95% CI: 0.02-0.05, P = 3 x 10^-4^).

### Associations of metabolomic age gap with risk factors and ageing phenotypes

Obesity, at risk of depression, and current use of statins were all found to be consistently associated with increased metabolomic age gap in our main model (with nominal P < 0.05 in both cohort analyses). Additionally, although only reaching statistical significance in one of the two cohorts, age gap estimates were found to be higher for male sex (Airwave), diabetes (TILDA), hypertension (Airwave), and physical inactivity (TILDA). No effects with age gaps (P>0.05) were observed for alcohol use, current smoking, education attainment, or household income in either cohort (Figure 6A). We also examined the relationship of metabolomic age gap models with age-related phenotypes available in TILDA. Higher levels of glycated haemoglobin HbA1c (biomarker of fasting glucose), C-reactive protein (inflammation), polypharmacy, timed up and go test (mobility), choice reaction time tasks (cognitive function), and lower estimated glomerular filtration rate (eGFR, kidney function) were associated with increased metabolomic age gap in the cross-sectional analysis (Figure 6B).

**Figure 6.**
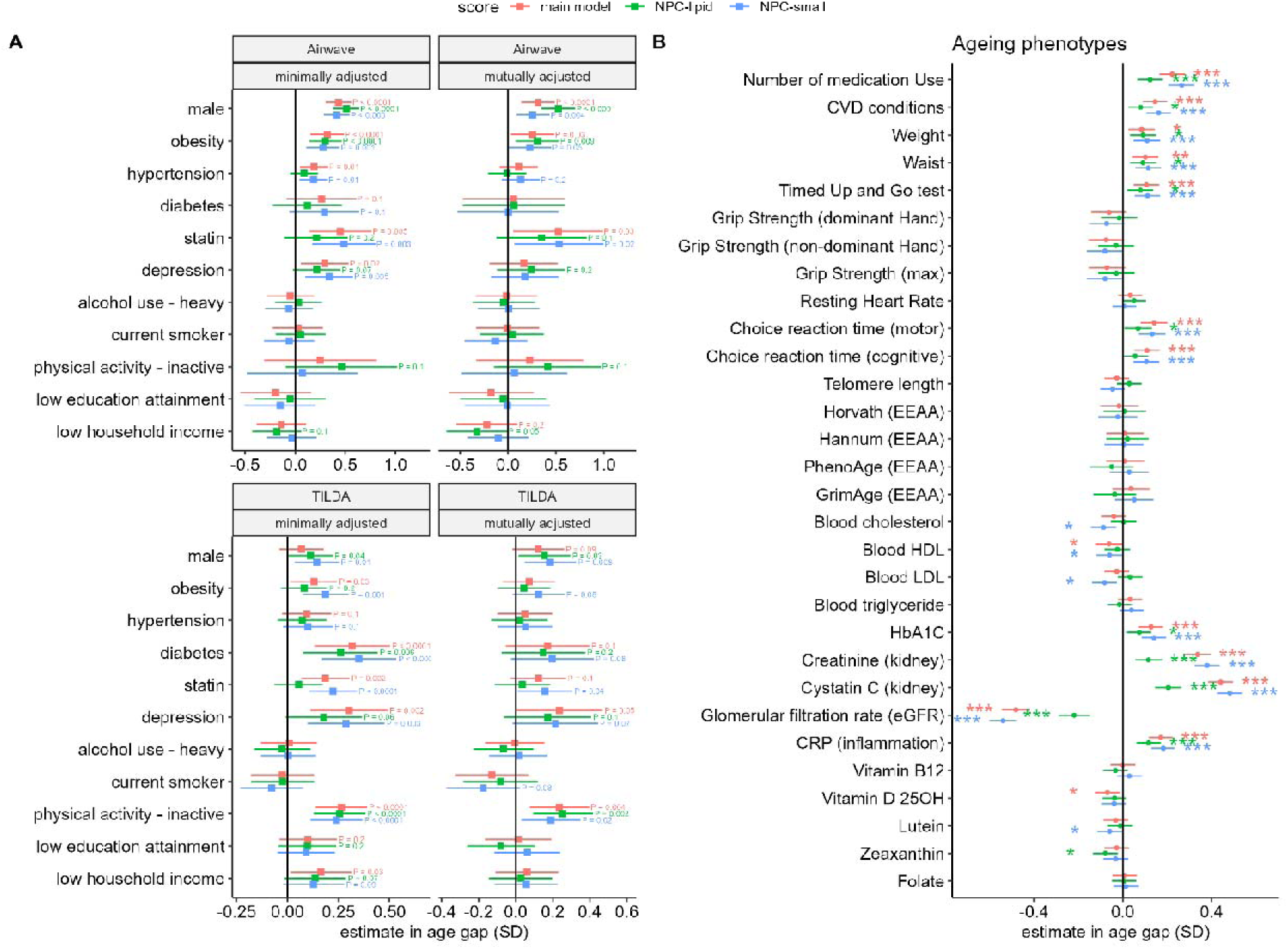
Metabolomic age gap with risk factors and ageing phenotypes. Age gaps were standardized and treated as dependent variable in linear regression models adjusted for age and sex, except when examining age gap associations with sex (A) Risk factor associations in Airwave and in TILDA. (B) Associations of Metabolomic age gap and ageing phenotypes in TILDA. * denotes P < 0.05, ** denotes P < 0.001; *** denotes associations significant after accounting for multiple testing using Bonferroni correction.

Within approximately 400 and 900 TILDA samples with epigenetic age and telomere length data, respectively, we found no associations with either epigenetic age acceleration or telomere length, suggesting the metabolomic age gap is complementary to other biological age indicators.

## Discussion

Through the application of broad-spectrum non-targeted mass spectrometry in two large national populations spanning the full adult lifespan from the UK and Ireland, we have characterised the profound relationship between ageing and the blood metabolome. We have further identified shared metabolic changes between ageing, frailty, and mortality risk, providing mechanistic insight into age-related vulnerability. We present a new metabolomic “clock” to assess overall metabolic ageing that combines high predictive performance of chronological age, interpretability and broad pathway coverage. We find that the metabolomic age gap derived from this clock appears to be a powerful measure of biological ageing, as it is associated with key ageing phenotypes including cognitive impairment and mortality, providing additional predictive value of 10-year mortality risk over standard clinical risk factors. This highlights its potential for applications such as improved prognostic risk stratification in population settings or as an endpoint in anti-aging trials.

Specifically, one standard deviation increase in metabolic ageing was associated with a 43% higher risk for mortality. This effect size is similar to that reported for the metabolomic clock of Sebastiani et.al ^35^, those reported in proteomic clocks studies ^36^ ^37^, and much stronger than those of epigenetic clocks similarly trained on chronological age ^38^. We observed that accelerated metabolomic aging at baseline was associated with functional aging measures including the Fried frailty index, MCI assessed through MoCA, and with poorer cognitive function assessed longitudinally through the MMSE assessments. While several metabolites have previously been identified as early predictors for MCI ^39^ ^40^ ^41^ ^42^, this is the first study to our knowledge that demonstrates an association between metabolic ageing and MCI.

The importance of metabolic factors is increasingly recognised in the pathogenesis of dementia ^43^, particularly mediated through mitochondrial dysfunction, a primary hallmark of ageing. Multiple acylcarnitine species, which are key to transport and beta-oxidation of fatty acids within the mitochondria ^44^, were among the most important contributors to the metabolomic clock, underlining the importance of mitochondrial function to metabolic aging and potentially dementia pathology. We also found the metabolomic age gap to be associated with increased risk of depression in both TILDA and Airwave. Whilst a few studies have tried identifying metabolic signatures of depression ^45–47^, except for our own earlier study involving Airwave ^9^, no studies have found a significant association between higher depression risk and increased metabolomic age gap. Two large studies applying NMR-based metabolic age assessments ^48^ ^49^ observed no association with depression, which may be attributed to the more comprehensive metabolic ageing assessment provided by LC-MS.

The ensemble LASSO learning approach employed here improved cross-sectional model prediction accuracies, and the predicted metabolomic age of our main model had a correlation coefficient *R* > 0.9 and a mean absolute error of 4.8 years in the test data. This is consistently better than chronological ageing models trained using NMR-based metabolomics datasets, which have *R* < 0.7, and MAE > 5 years in the UK Biobank samples ^12^, and comparable to age prediction performance among the best performing LC-MS-based metabolomic ageing metrics ^9,14,50^. This is particularly impressive considering that the ageing clock was trained using two cohort samples with distinct population characteristics. For instance, using metabolomics data acquired through Metabolon, Sebastiani et.al ^35^ demonstrated only relatively modest accuracy (MAE ∼ 7 years) of chronological age prediction with their metabolomic clock in the repeated samples of the Long Life Family Study. Although our data suggest that the model-predicted age is consistently higher at follow-up visit than at baseline, the mean Δ predicted age is less than the study follow-up interval (Δ calendar age) in both our study cohorts, implying that metabolomic age currently lacks accuracy in predicting changes in participants’ chronological age in longitudinal study settings. However, we are not aware of any metabolomic age study that explicitly evaluates their model age prediction longitudinally, and we speculate that this is an inherent limitation of age models derived from cross-sectional data, which manifests as regression toward the mean ^51^. Furthermore, there is currently little information in the literature on whether measurements of metabolomic age gap are reproducible between study visits from the same population, and our findings that age gaps were stable across the two visits (Pearson’s r > 0.6) demonstrates that metabolomic age gaps convey participant-specific health information.

The most frequently selected metabolites in our main multivariable ageing model represent diverse metabolic pathways and routes of exposure, and they included steroid sulphates and glucuronides, as well as acyl carnitines (beta-oxidation), phenylacetylglutamine (gut microbial metabolite from digestion of dietary proteins in plants or animals ^52,53^), niacinamide (vitamin B3), 4-hydroxychlorothalonil (fungicide metabolite), gamma-Glu-cystine (precursor to antioxidant glutathione), and MHPG-S (sulphated metabolite of neurotransmitter norepinephrine). Likewise, phenylacetylglutamine is a common chronic kidney disease toxin found associated with the incidence of cardiovascular disease ^54^ and has been suggested to be causally related to platelet response ^55^. Steroid hormones are known predictors of biological age ^56^, and are likely reflective of age-related changes in the hypothalamic-pituitary-adrenal functions ^57^. Sulphated hormones are considered to be an inactive reservoir of hormones, and a decrease in DHEA-S levels has been linked to sarcopenia or Alzheimer’s disease (AD) ^58^.

Some endogenous metabolites that are strongly associated with chronological age may have little impact on mortality risk if they are inactive in circulation. Conversely, due to a combination of changes in endogenous factors such as gut absorption, metabolic efficiency^59^, and age-related behavioural changes, some dietary metabolites, such as essential nutrients and antioxidants, appear important indicators of the ageing process and could be highly relevant for protecting against disease development. For example, loss of appetite is common as one gets older ^60^, so some environmental factors may be considered natural consequences of normal ageing processes. In this study, we have implicitly treated dietary influence as an inseparable part of the normal metabolomic ageing process, rather than as a confounding factor. The potential to capture age-related changes across the continuum of endogenous and exogenous chemical exposures, in addition to processes traditionally considered part of biological ageing, represents a key distinction of metabolomics, over other omics data types.

In metabolome-wide association analyses, we found many circulating metabolites to be associated with chronological age in both populations, and the most significant age associations were derived from our small molecule profiling assays. Strongest associations of chronological age were mainly glucuronides, sulphated, and steroid hormone (inversely associated) compounds and metabolites including C-glycosyl-tryptophan, the furan fatty acid CMPF, and acylcarnitines, replicating previous reports^8^. However, we identified several age-related metabolites that were previously poorly characterised in ageing epidemiology research, such as sialyllactose, the furan fatty acid 3-CMPFP and MHPG-S. Utilising the Irish TILDA sample, we examined and compared metabolic associations with all-cause mortality and frailty against chronological age associations. Furan fatty acids were unrelated to these outcomes suggesting they represent neutral or potentially adaptive age-related changes^61^.

However, both sialyllactose and MHPG-S were strongly related to mortality risk indicating they may represent pathways of interest for understanding age-related disease risk. Although little is known about the role of sialyllactose in blood, it is an oligosaccharide derived from N-acylneuraminic acid and likely has a signalling role through glycation of macromolecules. For instance, GlycA, a compound NMR based biomarker derived largely from N-acylneuraminic acid side chains on glycoproteins ^62^, is established as an important marker of systemic inflammation ^63^. MHPG-S is critical in norepinephrine metabolism, particularly in the brain, and has been used as an indicator of CNS noradrenergic activity in studies of depression and dementia ^64,65^.

The most significant associations of age-adjusted frailty were PUFA-containing triglycerides, carotenoids, and vitamin A1. Few large epidemiological studies have examined the metabolic associations with frailty. An early influential study ^66^ of frailty suggested that oxidative stress resulting from diminished antioxidant levels could be a vital pathway in the pathogenesis of frailty ^66^. We have found six metabolites detected to be simultaneously associated with chronological age, frailty, and mortality risk, including C-glycosyltrytophan, a bile alcohol glucuronide, N_2_,N_2_-dimethylguanosine and a diet-derived carotenoid. C-glycosyl tryptophan has previously been associated with CKD incidence ^67^, and multimorbidity^68^. Importantly, N_2_,N_2_-dimethylguanosine has previously been shown to be dependent on enzymatic activities of tRNA methyltransferase 1 (TRMT1), its deficiency may increase oxidative stress^69^, and higher level of N_2_,N_2_-dimethylguanosine has previously been found to be associated with increased mortality risk ^35,70,71^, higher incidences of CKD, cancer diseases ^72–74^ and hypertension ^75^.

Metabolomic age gaps were generally not sensitive to smoking and alcohol use. The relationship between smoking status and epigenetic age acceleration has previously been investigated, but other than GrimAge, which incorporated smoking pack-years into its training, the reported associations are generally either very weak or non-significant ^76,77^. There is even less evidence for the influence of smoking using proteomics/metabolomics-generated biological ageing metrics. Moreover, it also appears that alcohol use is not strongly associated with biological age measured by most epigenetic clocks ^78^. Metabolomic age gaps were not found to be associated with epigenetic age gaps or telomere length. In a multi-omics study of mental health, Jansen *et al.* ^49^ built their own telomere, epigenetic, proteomic, and metabolomics clocks within a single cohort and nonetheless, found only relatively weak correlations between epigenetic and transcriptomic clock age gaps (r = 0.15), and between proteomic and metabolomic clock age gaps (r = 0.19). This is consistent with our previously reported results in Airwave ^9^, reaffirming that epigenetics or telomere length and metabolomics inherently capture complementary and yet largely distinct ageing processes ^6,79^.

Limitations of our work include the use of non-fasted blood samples, since transient postprandial effects may have introduced noise and obscured the longer-term metabolic signals from ageing or health conditions. Our study samples are overwhelmingly of White European descent, which may have a different disease risk profile compared to other, less studied populations. Although we included both independent test samples and longitudinal evaluation for validation, we have not yet replicated our model results in an independent study cohort. Key strengths of our study include the inclusion of two large independent national samples spanning the full adult lifespan in our model training and analyses, the additional metabolite annotation work performed, validation of chronological age-metabolite associations across metabolomic platforms, and the longitudinal assessment of mortality risk and cognitive function in older adults. Importantly, the use of long-term reference samples in our metabolomic acquisitions has allowed harmonisation across study datasets and enhances the applicability of our metabolomic clock for future ageing studies. Further work should focus on validation in diverse populations and multi-omic integration for further mechanistic and causal insights.

In summary, the metabolomic clock developed and validated here appears to be an informative and reproducible marker of generalised age-related disease risk. Ultimately, it may provide complementary information relevant to personalised prevention and intervention strategies, beyond traditional clinical markers.

## Data Availability

The data supporting the findings of this study are available upon application to the Data Access Committee for the Airwave cohort (https://police-health.org.uk/applying-access-resource). TILDA data is deposited in the Irish Social Sciences Data Archive (ISSDA) https://www.ucd.ie/issda/data/tilda/

## Acknowledgement

This study was supported by the UK Research and Innovation Future Leaders Fellowship (MR/S03532X/1). This work at the National Phenome Centre was supported by the Medical Research Council and National Institute for Health Research (Grant number MC_PC_12025), and infrastructure support was provided by the National Institute for Health Research (NIHR) Imperial Biomedical Research Centre (BRC). The TILDA study was supported by funding from the Department of Health (TILDA-2017-1), Science Foundation Ireland (SFI)/Research Ireland and the Health Research Board (HRB) (SFI-19/US/3615) and an Irish Research Council Advanced Laureate Award (IRCLA/2023/2029). The Airwave Health Monitoring Study was funded by the UK Home Office (780-TETRA, 2003-18) and the Medical Research Council (MR/R023484/1, 2018-23) during its initial phase and is currently funded by UKRI grant award UKRI2539. The study acknowledges support from the National Institute for Health and Care Research (NIHR) Health Protection Research Unit (HPRU) in Radiation Threats and Hazards (NIHR207424) with additional infrastructure support provided by the NIHR Imperial College Biomedical Research Centre. The views expressed are those of the author(s) and not necessarily those of the NIHR, UK Health Security Agency or the Department of Health and Social Care. PE is a UK DRI Professor at the UK Dementia Research Institute, Imperial College London (funders include the Medical Research Council, Alzheimer’s Society and Alzheimer’s Research UK).

## Author Contributions

OR conceived and supervised the study. CL performed the data analysis and drafted the initial manuscript. EC conducted metabolite feature annotation and identification. RP assisted with data visualisation. AO and DC managed the TILDA biobank and were responsible for sample selection, retrieval and plating. AD, IT, PE, RAK, CM contributed to cohort study design and management. All authors have contributed to the review and editing of the manuscript.

## Abbreviation

BMI: body mass index
LPC: lysophosphatidylcholine
PC: phosphatidylcholine
LCMS: liquid chromatography-mass spectrometry
NMR: nuclear magnetic resonance
TILDA: Irish Longitudinal Study of Aging
PUFA: polyunsaturated fatty acid
DHEA-S: dehydroepiandrosterone sulfate
CI: confidence interval
SD: standard deviation
MoCA: Montreal Cognitive Assessment
MMSE: Mini Mental State Examination
TILDA: The Irish Longitudinal Study of Aging
Airwave: the Airwave Health Monitoring Study
WRAP: Wisconsin Registry for Alzheimer’s Prevention
VETSA: Vietnam Era Twin Study of Aging

---

**Figure S1.**
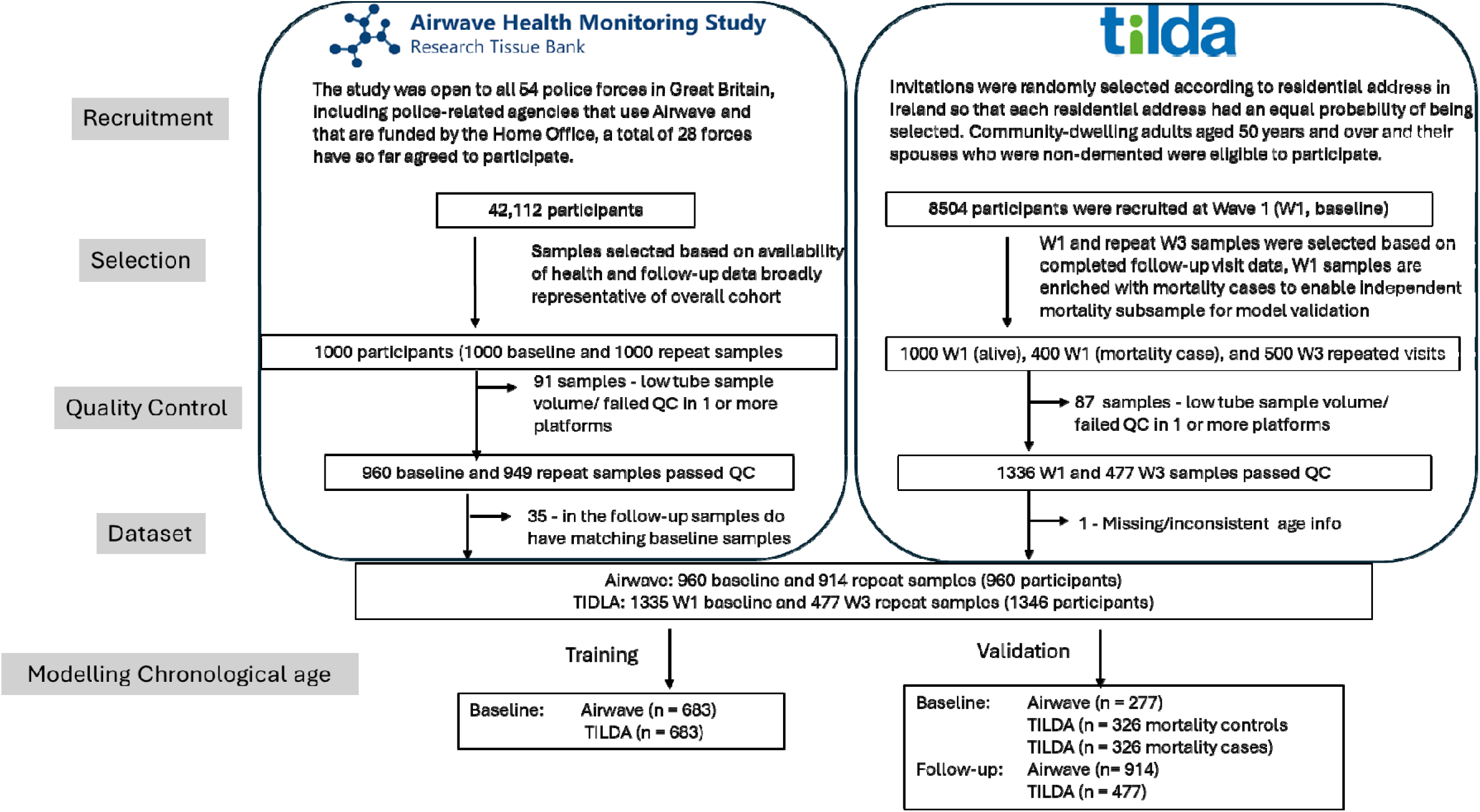
Summary of participants included in the study

**Figure S2.**
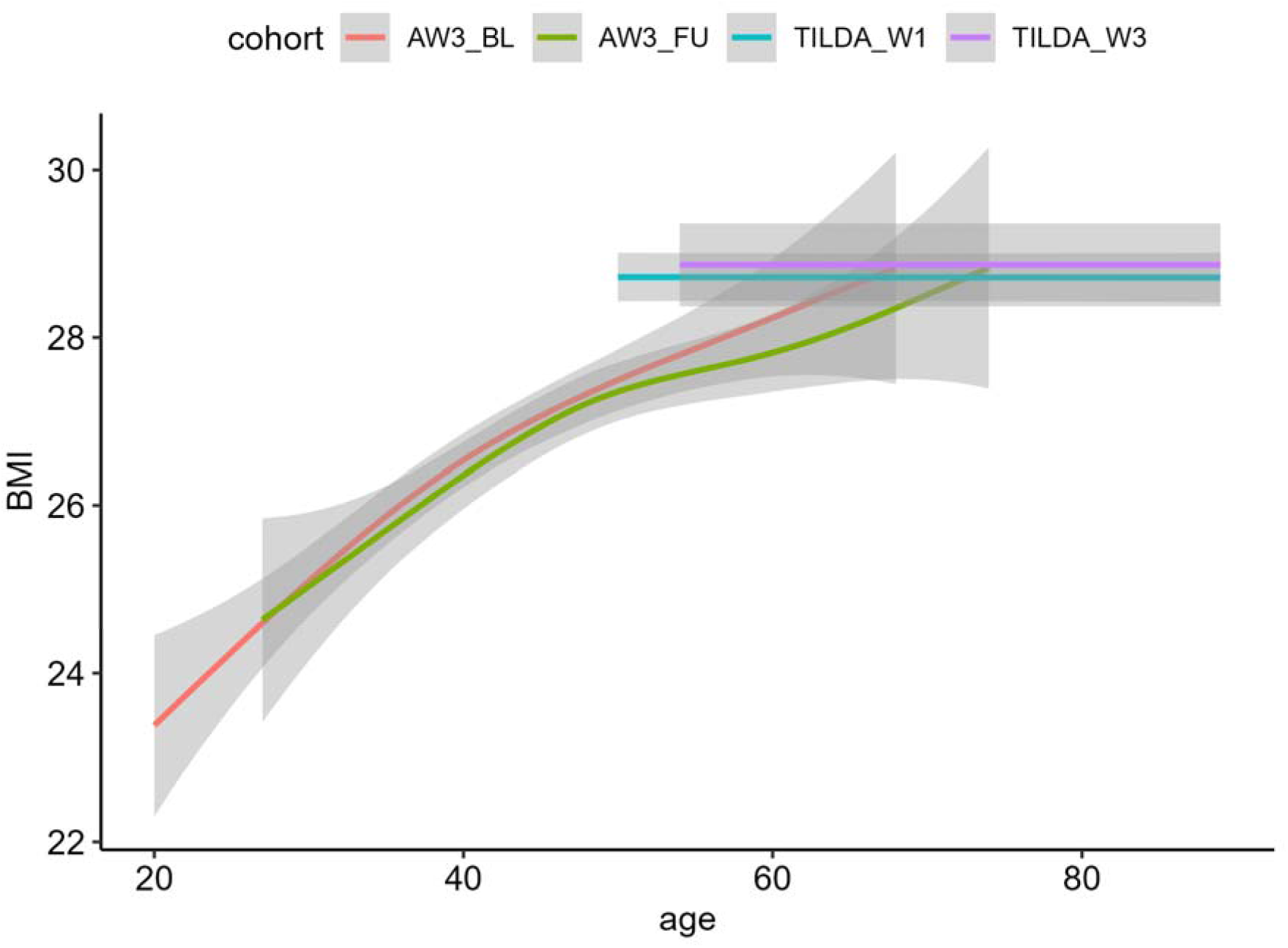
Relationship between age and body mass index in Airwave and TILDA

**Figure S3.**
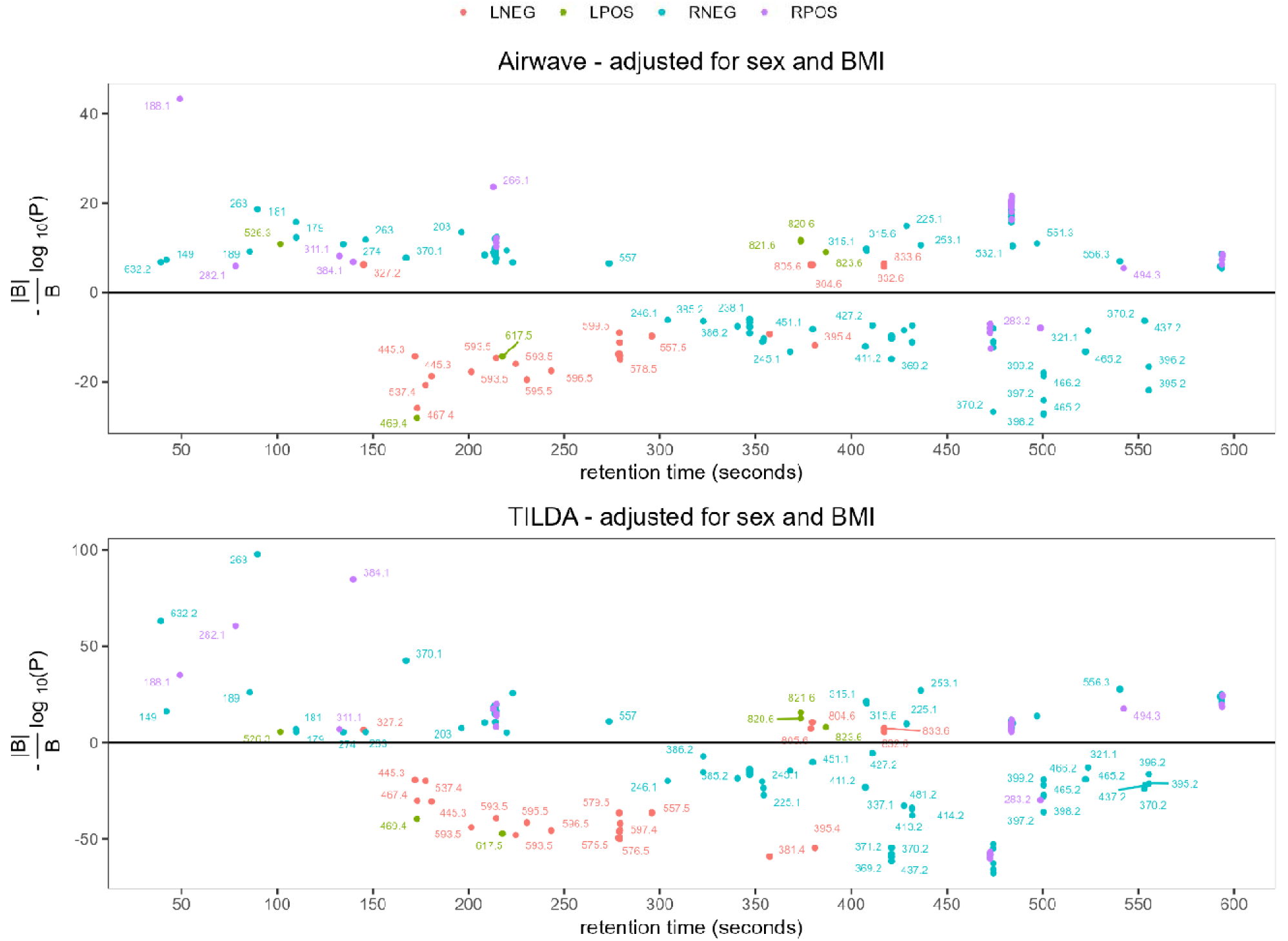
Metabolomic wide association study of chronological age in Airwave (top) and TILDA (bottom). The plot highlights the 173 XCMS mass spectrometric features that are concordantly associated with chronological age in both study cohorts in the four mass spectrometry platforms (LPOS, LNEG, RPOS, and RNEG), and the feature ions are labelled according to their *m/z* values.

**Figure S4.**
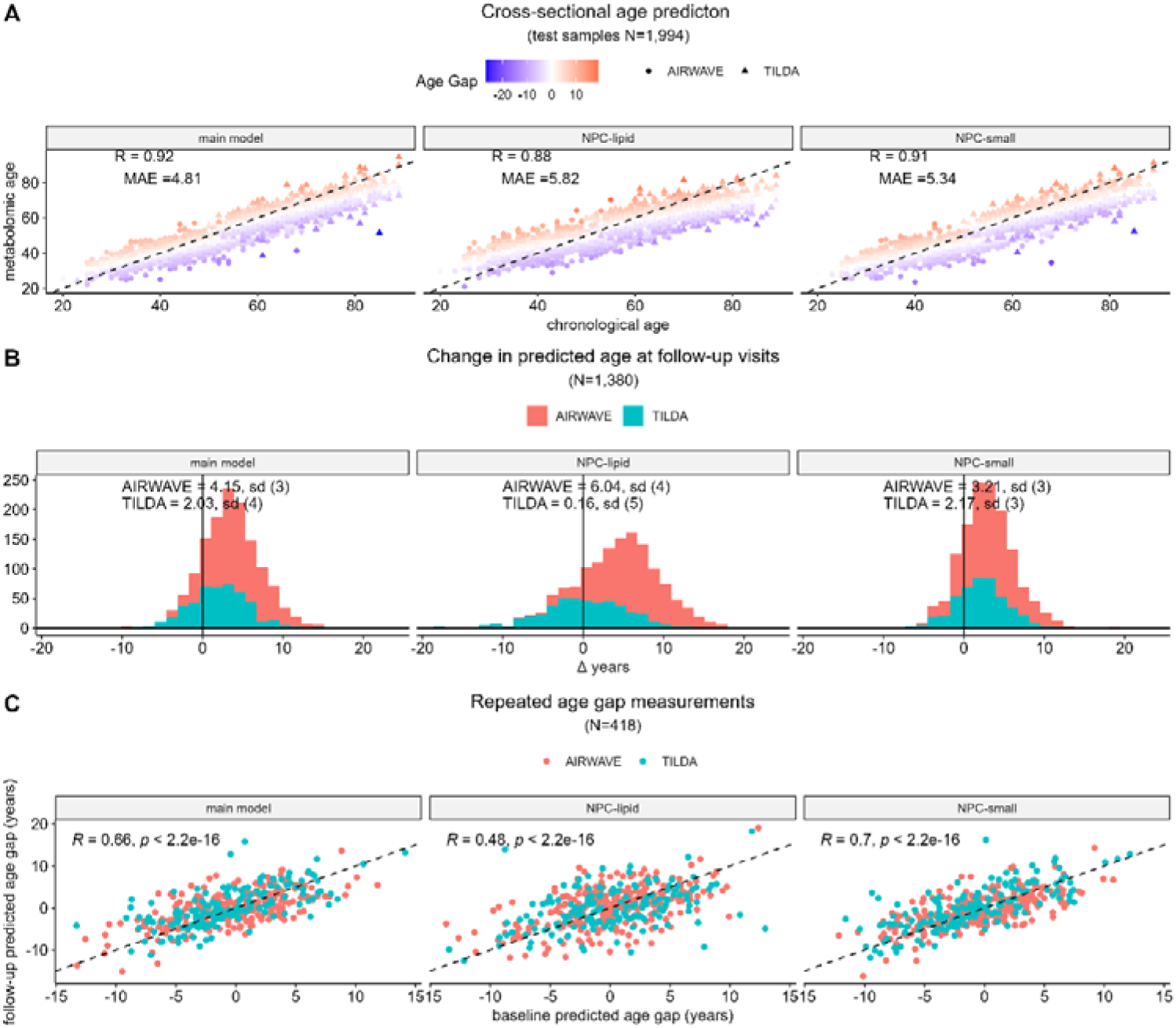
Age prediction performance for the three chronological ageing clocks – main model, NPC-lipid model and NPC-small molecule model. A) Cross-sectional performance in the test samples B) Changes in predicted age at follow-up visits C) Scatter plot of metabolomic age gaps independently estimated at baseline (x-axis) and follow-up visits.

**Figure S5.**
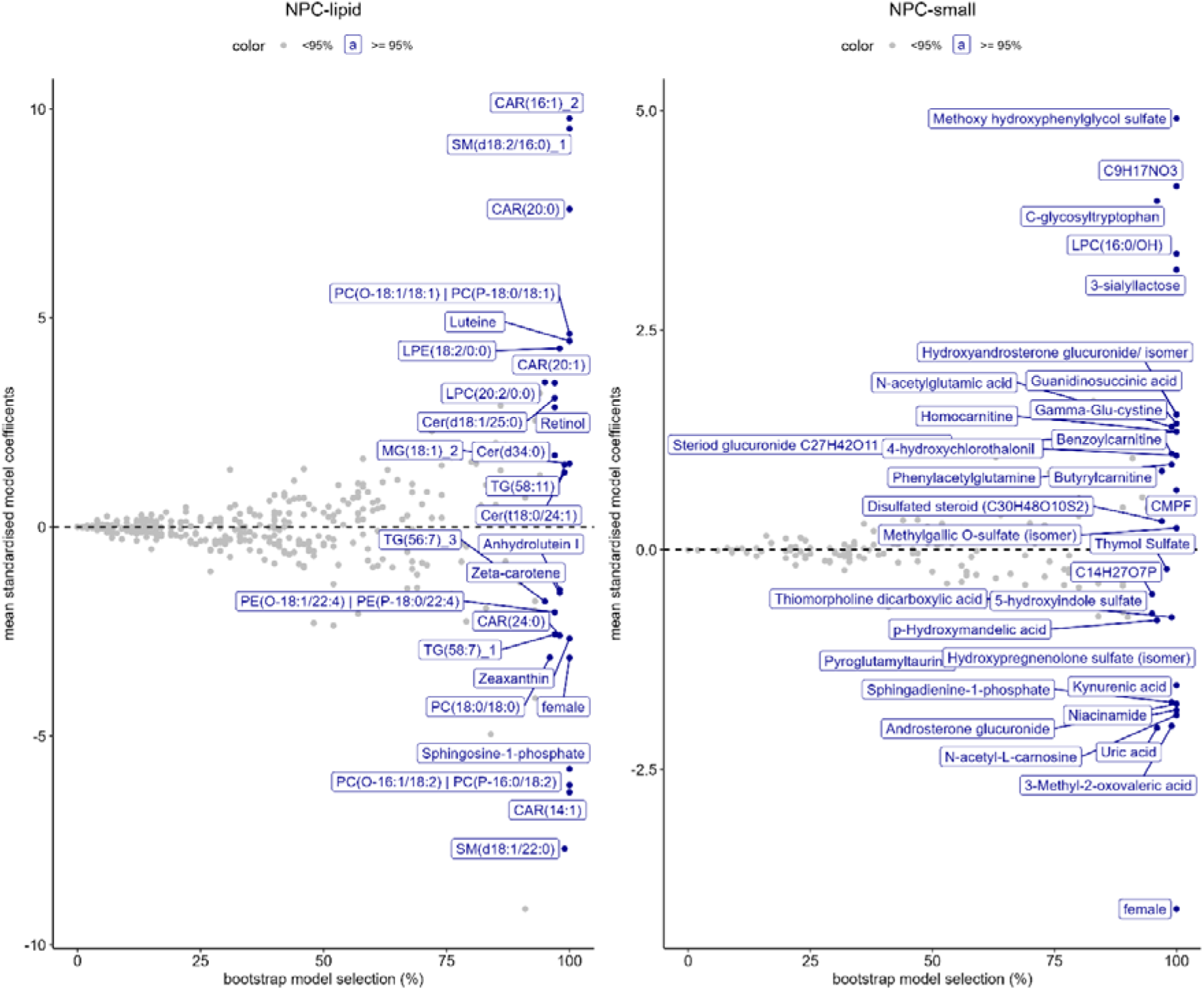
Bootstrap aggregate LASSO model of chronological age. Metabolites selected in ≥ 95% of the bootstrap models are highlighted in blue.

**Figure S6.**
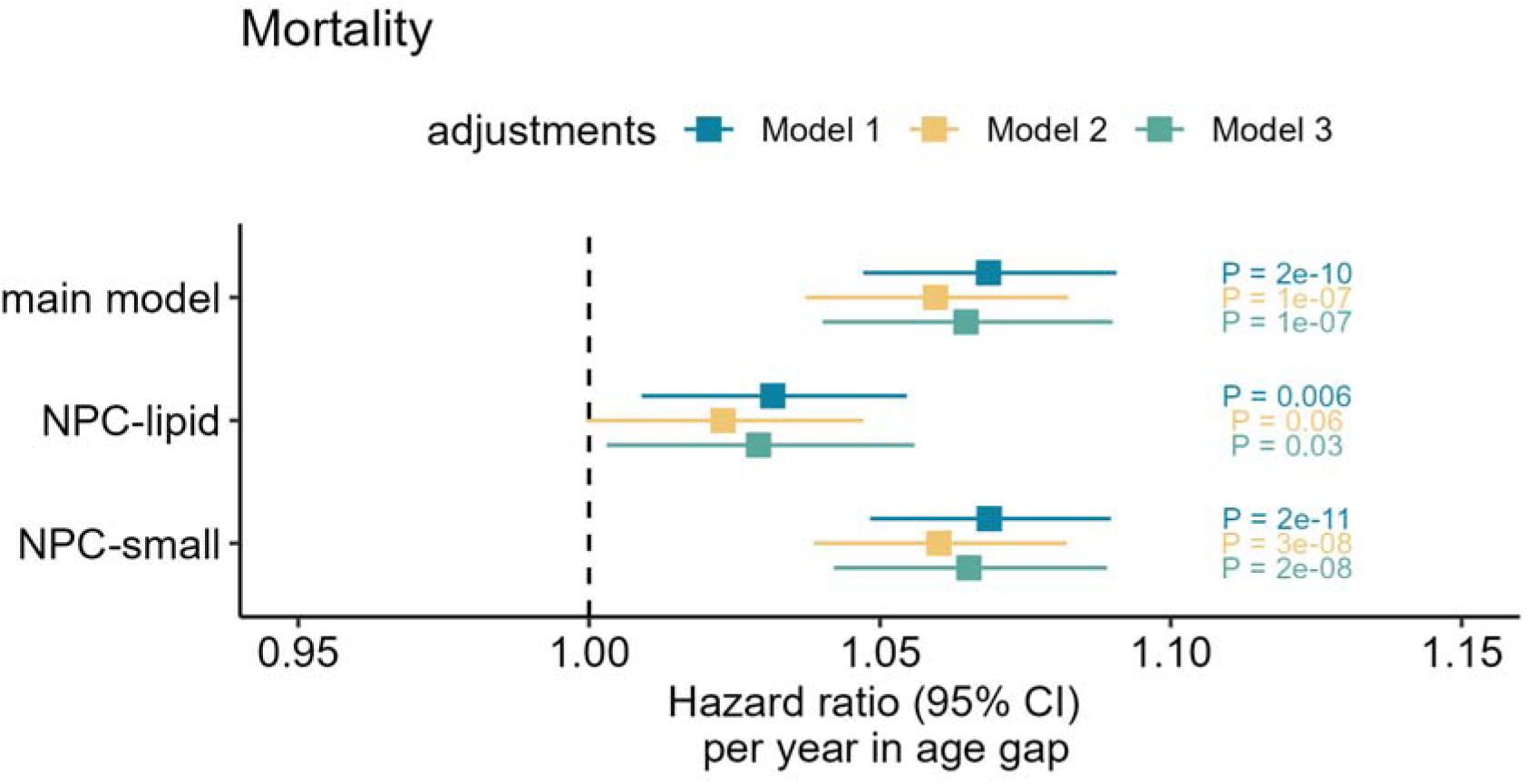
Associations between age gap (expressed in years) and all-cause mortality, in TILDA controls and mortality case samples matched 1:1 for age, sex and BMI (N case = 326) excluded from model training. Model 1adjusted for age and sex, Model 2 additionally adjusted for BMI, hypertension, diabetes, statin use, number of medications used, Model 3 additionally adjusted for depression, education, smoking, alcohol use and physical activity.

**Figure S7.**
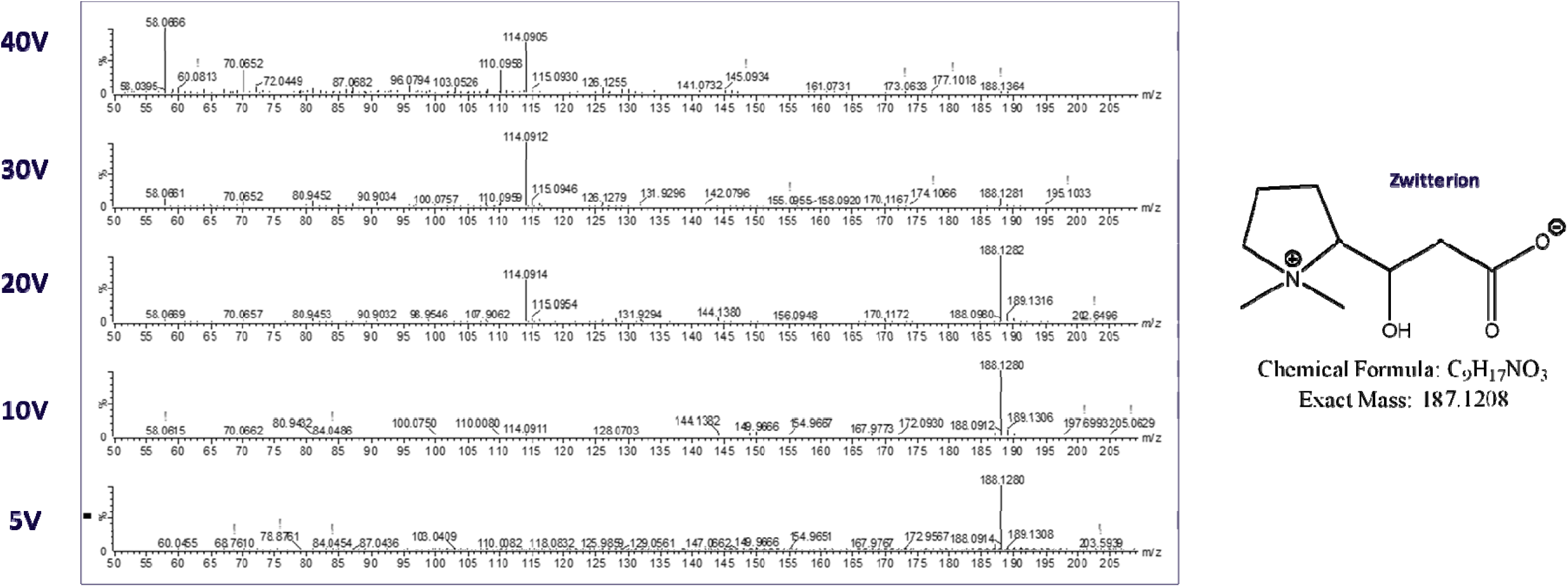
Putative Annotation of C_9_H_17_NO_3_. Q-TOF MS/MS at different collision energy levels in the positive ionization mode and suggested putative structure in the neutral – zwitterionic form. Owing to its significance, we carried out additional MS/MS invest gation and an extensive spectral interpretation effort along with database searches to suggest a possible structure of this unknown molecule. The MS/MS analysis of the parent ion was performed at a range of increasing collision energies, making it possible to follow the appearance and intensity of different detected fragments, some of which were key for structural elucidation. For instance, the fragment at m/z 70.065 is characteristic of the presence of proline moiety while the fragment at m/z 58.065 is specific for betaines of amino acids (https://doi.org/10.1002/jms.2029). Since the loss of C3H9N (59 Da) was not detected in the MS/MS of C9H17NO3, we suggest that the structure contains a proline betaine moiety. In addition, the fragment at m/z 170.118 corresponds to the loss of a water molecule from the parent ion, indicative of the possible presence of an hydroxyl group in the structure while the fragment at m/z 144.139 points to a loss of CO2 and likely presence of a carboxylate group in the parent molecule. The retention time of the unknown molecule in the reversed-phase assay in positive ionization mode is < 1 min, and it elutes among other polar molecules such as acylcarnitines (for instance, L-carnitine, delta-valerobetaine, acetylcarnitine, proline betaine, etc.) which proves its polar nature and structural similarity with known molecules containing a betaine moiety. The structure proposed for this unknown has some similarity with the structure of homocarnitine, reported by Weinberg, et al. (doi: 10.1016/j.jbc.2024.108074) and found to be associated with age in this work. The molecular formula of homocarnitine is C8H17NO3, which has only one less carbon atom C compared to the unknown molecule, which can be only possible with the cyclic structure of the unknown molecule.

